# Clonal Hematopoiesis is Associated with Risk of Incident, Seropositive Rheumatoid Arthritis

**DOI:** 10.1101/2024.09.20.24314099

**Authors:** Kun Zhao, Yash Pershad, J. Brett Heimlich, Michelle Ormseth, C. Michael Stein, Brian Sharber, Caitlyn Vlasschaert, Alexander G. Bick, Robert W. Corty

## Abstract

**Objectives:** Clonal hematopoiesis (CH), defined by acquired mutations in hematopoietic stem cells, has been linked to many inflammatory diseases of aging. We investigated whether CH subtypes such as clonal hematopoiesis of indeterminate potential (CHIP) and mosaic chromosomal alteration (mCA) are associated with the risk of incident rheumatoid arthritis (RA) and whether complement modifies these associations.

**Methods:** CHIP was ascertained in NIH AllOfUs, Vanderbilt BioVU, and the UKBiobank; mCA was detected in UKBiobank. A consistent phenotyping algorithm was applied across biobanks to identify seropositive RA (SPRA) and seronegative RA (SNRA). Age-scale survival models assessed the effect of CH on risk of incident RA. Effect modification was tested with interaction models with genetically predicted complement protein levels.

**Results:** Among 612,989 eligible participants, 30,840 had CHIP, 2,110 had incident SPRA, and 1,310 had incident SNRA. CHIP was associated with an increased risk of incident SPRA (HR 1.29; CI 1.05-1.57; *p*=1.3×10^-2^) driven primarily by *DNMT3A*-mutated CHIP (HR 1.42, CI 1.1-1.82, *p*=7.1×10^-3^) but no risk of SNRA. In UKBiobank, autosomal mCA and mosaic loss of Y (mLOY) were associated with an increased risk of incident SPRA (HR 2.12 and 2.85; CI 1.18-3.8 and 1.57-5.19; *p*=1.2×10^-2^ and 6.1×10^-4^), but no risk of SNRA. Higher genetically predicted levels of C1r and C1s attenuated the CHIP-SPRA association.

**Conclusions:** *DNMT3A*-CHIP, autosomal mCA and mLOY are novel risk factors for incident SPRA but not SNRA, supporting a serostatus-specific link between somatic mutation in epigenetic regulators and RA, with the classical complement pathway as a potential modifier of this risk.

## Introduction

Rheumatoid arthritis (RA) is a systemic, autoimmune disease characterized by chronic inflammation and heterogeneous clinical trajectories across dimensions of age, serologic status, and treatment response.^1^ RA primarily affects older adults, though people of all ages can be affected.^2^ RA diagnosed in a person of age 60 or more has been termed late-onset RA (LORA).^3^ LORA has a distinct genetic profile, higher levels of inflammatory cytokines, and inferior response to therapy.^4,5^ The mechanisms driving LORA remain incompletely understood. There is a pressing need for advancements in the diagnosis and care of LORA as epidemiologic data demonstrate a marked and continued rise in its incidence.^2^ The pathogenesis of seropositive RA (SPRA; defined by presence of rheumatoid factor and/or anti-cyclic citrullinated peptide antibodies in the serum) is known to involve antigen-driven clonal selection on lymphocytes.^6^ But the role of clonal selection among hematopoietic stem cells, a common phenomenon in older adults, remains much less explored.

The two most common forms of clonal hematopoiesis (CH) are clonal hematopoiesis of indeterminate potential (CHIP) and mosaic chromosomal alteration (mCA). CHIP is defined by the clonal expansion of hematopoietic stem cells carrying a small somatic mutation in leukemia-associated genes.^7–9^ Approximately 75% of CHIP cases are driven by mutation in one of three epigenetic regulators, *DNMT3A*, *TET2*, and *ASXL1*, with the remainder distributed over a long tail of ∼70 genes.^10^ CHIP prevalence is highly age-dependent: it rarely observed among people in their forties and younger (<1%) but seen in ∼15% of individuals in their seventies.^10^ Although typically asymptomatic, CHIP skews hematopoiesis toward myelopoiesis, causes chronic inflammation, and confers elevated risks for myeloid neoplasms, cardiovascular disease, and all-cause mortality.^8,11^ mCA, the other form of CH, is defined by large-scale chromosomal gain, loss, or copy-neutral loss of heterozygosity, of the autosomes or sex chromosomes (i.e., mosaic loss of chromosome X, known as mLOX, and mosaic loss of chromosome Y, known as mLOY). mCA is also common among older adults, can be asymptomatic, and confers risk for a distinct, but less-well understood, set of diseases of aging.^12,13^

Recent investigations have begun to elucidate the relationship between the CH and RA. Hiitola et al. found that CHIP is associated with prevalent RA in genotype- and seropositivity-specific manner, noting that the rate of *DNMT3A* CHIP is elevated among patients with SPRA and the rate of *TET2* CHIP is elevated among patients with SNRA.^14^ But disease ascertainment was based solely on diagnosis codes, an approach that has been found to have specificity of around 55%.^15–17^ Similarly, Uchiyama et al. highlighted the critical importance of age at disease onset, reporting that mLOY in males is a risk factor for LORA but potentially protective against young-onset RA (YORA).^18^ However, both studies had limited access to longitudinal clinical data. Furthermore, studies to date have offered limited insight into the mechanisms that link CH and RA. As the classical complement pathway can be activated by innate inflammation and plays a prominent role in RA pathogenesis, it represents a promising avenue for mechanistic study of the developing relationship between CH and RA.

To address these questions, we performed a multi-ancestry meta-analysis across three large biobanks: the NIH *All of Us* Research Program (AoU), Vanderbilt’s BioVU, and the UK Biobank (UKB). We leveraged extensive electronic health records (EHR) and genetic sequencing data to study over 600,000 research participants. We employed established bioinformatic pipelines for CH detection, a rigorous RA phenotyping strategy, and longitudinal study design to: (1) identify high-confidence cases of SPRA and SNRA, (2) comprehensively characterize the effects of CHIP and mCA on risk of incident SPRA and SNRA, (3) dissect the effects of CHIP on risk for CVD and mortality among research participants with RA, and (4) test for effect modification by genetically predicted protein levels.

## Methods

### Patient and public involvement

De-identified data was obtained from three biobanks with linked electronic health records (EHR) and either whole exome or whole genome sequencing (WES/WGS): the NIH All Of Us (AoU), Vanderbilt’s BioVU and the UK Biobank (UKB).

Patients and the public were not involved in the design, conduct, or reporting of this project. Inclusion/exclusion criteria and RA disease ascertainment criteria were applied consistently across the biobanks.

#### Inclusion criteria

Research participants were included in the study only if they had (1) adequate genetic data for CHIP ascertainment, (2) adequate clinical data for RA disease ascertainment (2+ diagnosis codes spanning a year or more, 1+ laboratory value, and 1+ medication prescription), and (3) adequate engagement with the healthcare system that RA would likely be diagnosed if it were present (i.e., 5+ total diagnosis codes).

#### Exclusion criteria

Research participants were excluded if they had age < 40 years at the time of the blood sample used for DNA sequencing because CHIP is very rare among the young.^9,19^ Similarly, research participants were excluded if they had prevalent hematologic cancer (defined by diagnosis codes in **Supplementary Table 1**).^7^ As smoking is a risk factor for both CH and RA, smoking status was a critical covariate. It was inferred from questionnaires administered to all participants in AOU, BioVU, and UKB and participants who did not answer were excluded.

#### Follow-up

Electronic health records in AoU and BioVU are updated approximately annually to include all available data, thus follow-up is until last available diagnosis code. In UKB, ambulatory records are kept by general practitioners and are available for approximately 45% of the cohort with a truncation date in 2016 as described in the UKB data showcase, defining the censoring date for participants with no diagnosis of RA. Only participants with available primary care codings were included in the study **(Fig. 1)**.

**Figure 1.**
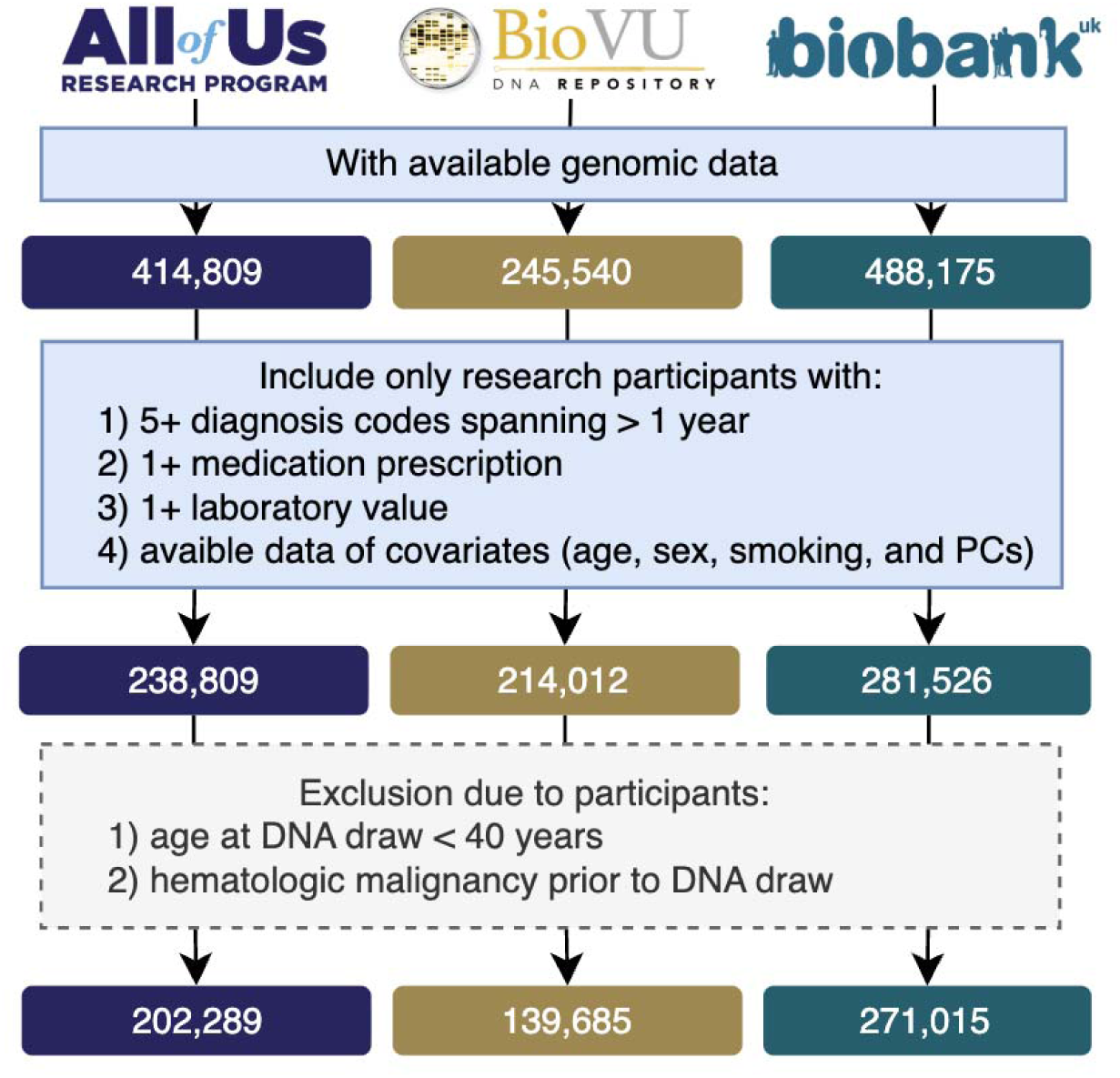
Study cohort flowchart across three biobanks. Flow diagram showing cohort selection in the *All of Us* Research Program (AoU), Vanderbilt BioVU, and UK Biobank (UKB). The final analytic cohorts included 202,289 participants from AoU, 139,685 from BioVU, and 271,015 from UKB. Many participants in AoU and UKB were not included due to lack of adequate electronic medical records data. Many participants in BioVU were excluded due to young age at blood draw.

### Rheumatoid arthritis ascertainment

We used a combination of diagnosis codes, laboratory data, and medication prescription to identify research participants with rheumatoid arthritis. This approach has been found to have sensitivity and specificity of approximately 85% and 99% respectively.^15,20–22^

#### Diagnosis codes

For AoU and BioVU, where diagnoses are recorded by ICD9 and ICD10 codes, codes matching ‘M05’ were considered as SPRA, codes matching ‘M06’ were considered as SNRA, and codes matching ‘714.0’ were considered as RA not otherwise specified (RA NOS). For UKB, where diagnoses are recorded in the SNOMED vocabulary, relevant codes were identified with a search strategy described in the **Supplementary Appendix**.

Two codes of a class (SPRA, SNRA, or RA NOS) were required to categorize the participant as that class. In cases where a participant was classified as both SPRA and SNRA, they were assigned to whichever class had a higher code count. In cases where a participant was assigned both a sero-specific class (SPRA or SNRA) and RA NOS, they were assigned to the sero-specific class.

#### Medications

The list of disease modifying anti-rheumatic drugs (DMARDs) was defined as the combination of conventional synthetic DMARDs, biologic DMARDs, and targeted synthetic DMARDs. For all three biobanks, DMARDs were identified by searching the OMOP ingredient table for values that match the regular expression that combines the list of DMARD names, then using the concept_relationship and concept_ancestor tables to find all drug prescriptions in the biobank that contain those ingredients.

#### Laboratory values

We identified concepts that capture rheumatoid factor (RF) and anti-cyclic citrullinated peptide (CCP) and developed a careful parsing scheme to assign each result as either positive or negative, as detailed in the **Supplementary Appendix**. Each participant was considered seropositive by laboratory values if they had a positive RF or CCP, negative if they had a result for at least one with no positive results, and N/A otherwise.

#### Case Definition

Patients were considered to have SPRA if they met either of these criteria:

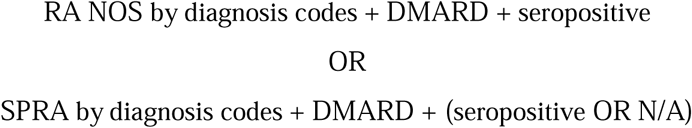

Patients were considered to have SNRA if they met either of these criteria:

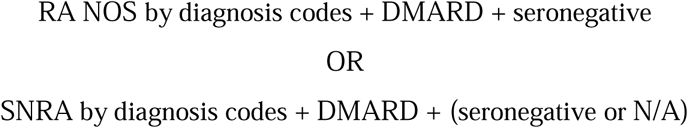

#### Date of disease onset

Participants with RA were considered to have developed the disease on the date the diagnosis code first appeared in their medical records. Cases were considered as “prevalent” when diagnosed on or before the date of the blood sample used for DNA sequencing and “incident” when diagnosed after the blood sample.

#### Osteoarthritis controls

Controls with osteoarthritis were identified in BioVU as participants with phecode MS_708 and no instances of M05 or M06.

### Clonal hematopoiesis of indeterminate potential ascertainment

Based on genetic sequencing data availability, CHIP calling was performed with WGS for AoU and BioVU and with WES for UKB. For each cohort, the same list of genes, variants, and analysis was used. Specifically, CHIP-defining somatic single nucleotide variants and short indels were identified per established variant detection methods as we have previously described.^25,26^ Briefly, 74 genes known to harbor CHIP-driving variants were screened for somatic variants using the *Mutect2* somatic variant caller from the GATK software suite to identify putative CHIP variants **(Supplementary Table 4)**.^8,27,28^ Putative CHIP variants were screened using the following criteria: total sequencing depth ≥ 20, alternate allele read depth count ≥ 3, representation in both sequencing directions, and statistical exclusion of germline heterozygosity.

### Mosaic chromosomal alteration detection

mCA detection in the UK Biobank was previously described. In brief, mCAs in blood DNA genotyping intensity data were detected using a validated statistical phasing-based approach.^29^ This method leverages long-range phase information to identify allelic imbalances, enhancing sensitivity for detecting large events at low cell fractions. mCAs were categorized into autosomal mCAs, mosaic loss of chromosome Y in males, and mosaic loss of chromosome X in females.

### Genetically predicted complement protein levels

To investigate the mechanistic role of the classical complement pathway without the confounding effects of active inflammation or reverse causation, we used genetically predicted protein levels as a proxy for stable, lifelong systemic exposure. Complement protein levels were estimated among participants of European ancestry in BioVU (N = 116,082) and UKB (N= 260,119).^28^ The single nucleotide variants and their weights are provided in **Supplementary Table 5**. The genetically predicted complement levels were calculated by summing the weighted genotypes of all selected variants and scaling to mean 0 and standard deviation 1.

### Statistical analyses

Descriptive statistics were used to summarize baseline characteristics. Continuous variables were reported as median (IQR), and categorical variables as counts and percentages.

The association between CHIP and incident RA was assessed using age-scale Cox proportional hazards models with covariates of sex, smoking status, and genetic principal components (PC1–PC5). Age of entry was defined as the age at which the blood sample was drawn that was used for DNA sequencing and age of exit as the age at diagnosis with RA (event) or the end of the available follow-up data (censoring). The proportional hazards assumption was evaluated with examination of Schoenfeld residuals. Logistic regression was used to test the effect of CHIP on risk for prevalent RA using the same covariates and additionally including age of blood draw, which is accounted for naturally in the survival analysis.

Inverse variance-weighted meta-analysis was performed under either a fixed- or random-effects framework, selected according to the degree of heterogeneity (I² statistic; **Fig. S1**).

Among participants with RA, the impact of CHIP on all-cause mortality was assessed using age-scale Cox proportional hazards models, adjusting sex, smoking status, and principal components of ancestry (PC1–PC5), as above. Kaplan–Meier curves were generated to visually compare survival across CHIP and non-CHIP groups, and differences were assessed using the log-rank test.

We tested for effect modification by modifying the age-scale survival analysis described above to include an interaction term between CHIP status and genetically predicted complement levels. All analyses were conducted by R version 4.2.0 (https://www.r-project.org).

## Results

### Sociodemographic characteristics

After applying the unified inclusion and exclusion criteria, a total of 612,989 participants were included across the three biobanks: 202,289 from AoU, 139,685 from BioVU, and 271,015 from UKB. Their sociodemographic characteristics are presented in **Table 1**.

**Table 1.**
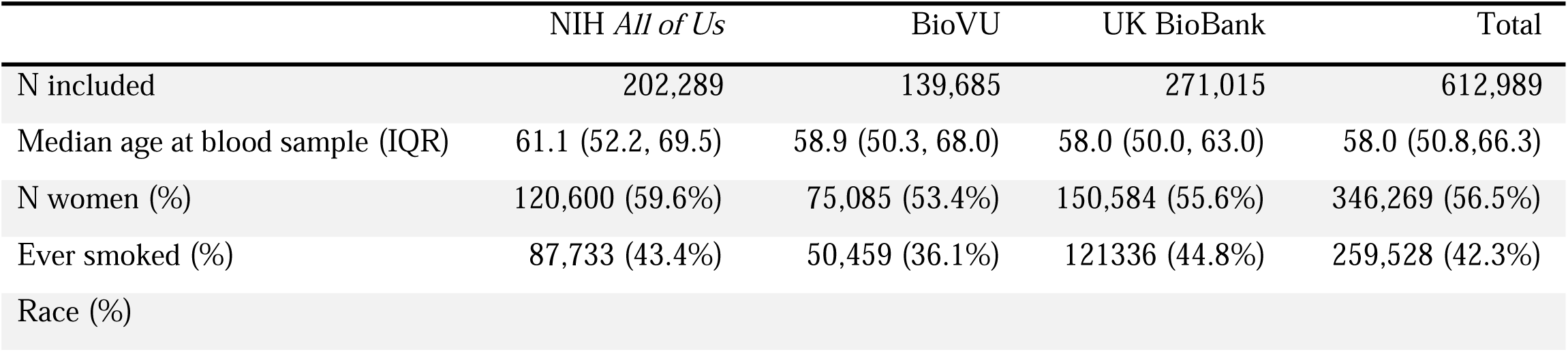
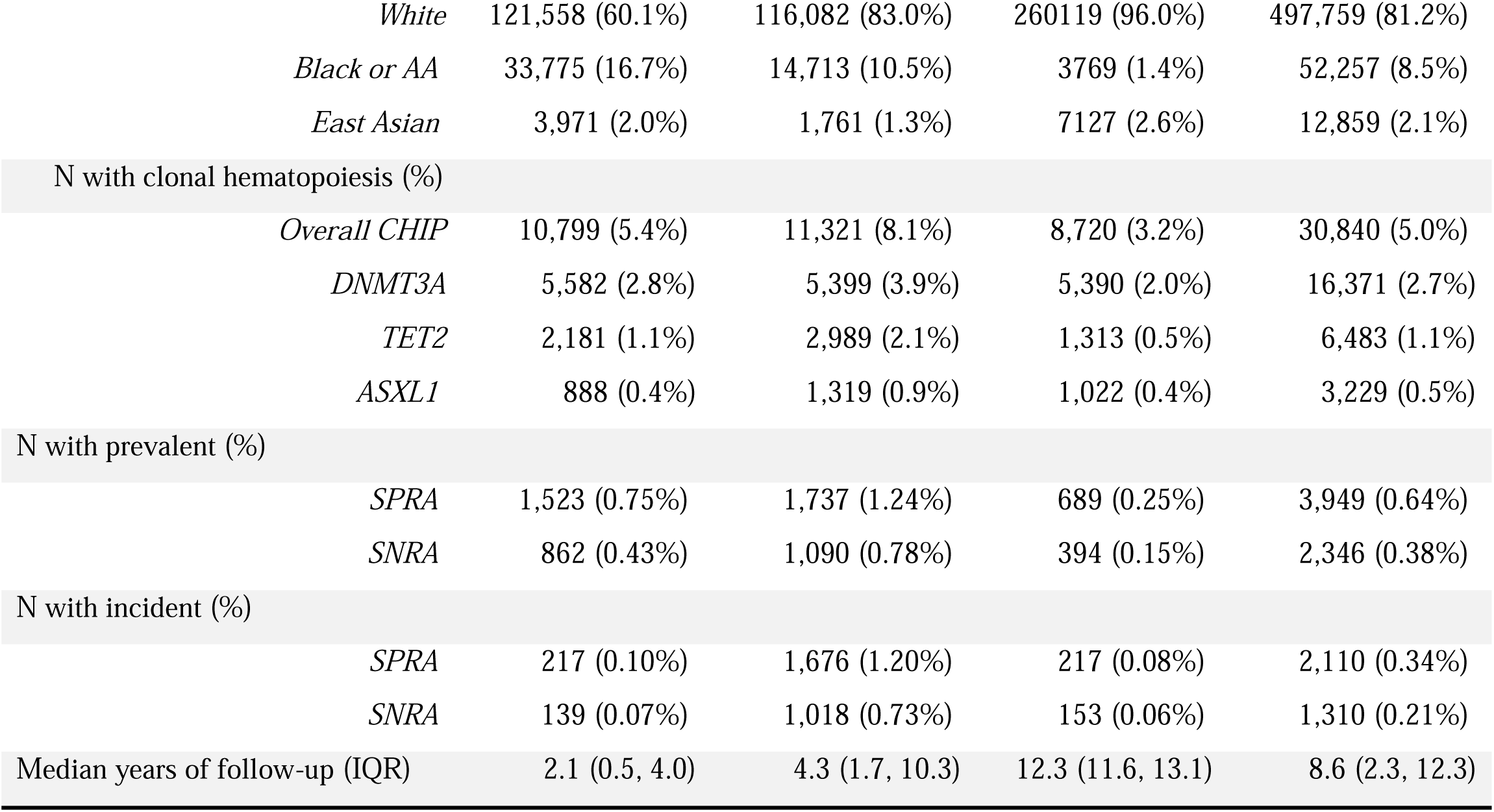
Socio-demographic characteristics of the three cohorts studied and in aggregate. .

|  | NIH *All of Us* | BioVU | UK BioBank | Total |
| --- | --- | --- | --- | --- |
| N included | 202,289 | 139,685 | 271,015 | 612,989 |
| Median age at blood sample (IQR) | 61.1 (52.2, 69.5) | 58.9 (50.3, 68.0) | 58.0 (50.0, 63.0) | 58.0 (50.8,66.3) |
| N women (%) | 120,600 (59.6%) | 75,085 (53.4%) | 150,584 (55.6%) | 346,269 (56.5%) |
| Ever smoked (%) | 87,733 (43.4%) | 50,459 (36.1%) | 121336 (44.8%) | 259,528 (42.3%) |
| Race (%) |  |  |  |  |
| *White* | 121,558 (60.1%) | 116,082 (83.0%) | 260119 (96.0%) | 497,759 (81.2%) |
| *Black or AA* | 33,775 (16.7%) | 14,713 (10.5%) | 3769 (1.4%) | 52,257 (8.5%) |
| *East Asian* | 3,971 (2.0%) | 1,761 (1.3%) | 7127 (2.6%) | 12,859 (2.1%) |
| N with clonal hematopoiesis (%) |  |  |  |  |
| *Overall CHIP* | 10,799 (5.4%) | 11,321 (8.1%) | 8,720 (3.2%) | 30,840 (5.0%) |
| *DNMT3A* | 5,582 (2.8%) | 5,399 (3.9%) | 5,390 (2.0%) | 16,371 (2.7%) |
| *TET2* | 2,181 (1.1%) | 2,989 (2.1%) | 1,313 (0.5%) | 6,483 (1.1%) |
| *ASXL1* | 888 (0.4%) | 1,319 (0.9%) | 1,022 (0.4%) | 3,229 (0.5%) |
| N with prevalent (%) |  |  |  |  |
| *SPRA* | 1,523 (0.75%) | 1,737 (1.24%) | 689 (0.25%) | 3,949 (0.64%) |
| *SNRA* | 862 (0.43%) | 1,090 (0.78%) | 394 (0.15%) | 2,346 (0.38%) |
| N with incident (%) |  |  |  |  |
| *SPRA* | 217 (0.10%) | 1,676 (1.20%) | 217 (0.08%) | 2,110 (0.34%) |
| *SNRA* | 139 (0.07%) | 1,018 (0.73%) | 153 (0.06%) | 1,310 (0.21%) |
| Median years of follow-up (IQR) | 2.1 (0.5, 4.0) | 4.3 (1.7, 10.3) | 12.3 (11.6, 13.1) | 8.6 (2.3, 12.3) |

### Clonal hematopoiesis of indeterminate potential

Across the three cohorts, a total of 30,840 (5.0%) participants had ≥1 detectable CHIP mutation(s), with rates of 5.4% in AoU, 8.1% in BioVU, and 3.2% in UKB. CHIP carriers were, on average, older than non-carriers (mean age 58.4 vs 65.8, *p*<0.001). Most CHIP clones were driven by mutations in *DNMT3A* (53%), *TET2* (19%), and *ASXL1* (9%), which together accounted for 82% of all detected driver mutations. All cohorts demonstrated a sharp increase in the rate of CHIP with age, but the absolute frequency differed across biobanks, with BioVU showing the highest prevalence at all age groups **(Fig. S2)**.

### Prevalent and incident rheumatoid arthritis

SPRA and SNRA were identified in 3,949 (0.64%) and 2,346 (0.38%) of the combined cohort, respectively. Incident SPRA (first diagnosed after the blood draw when CHIP status was ascertained) occurred in 217 (0.10%) participants in AoU, 1,676 (1.20%) in BioVU, and 217 (0.08%) in UKB, while incident SNRA occurred in 139 (0.07%), 1,018 (0.73%), and 153 (0.06%), respectively. Prevalent SPRA (first diagnosed before the blood draw when CHIP status was ascertained) occurred in 1,523 (0.75%), 1,737 (1.24%), and 689 (0.25%) of participants in AoU, BioVU, and UKB, respectively. Prevalent SNRA occurred in 862 (0.43%), 1,090 (0.78%), and 394 (0.15%) of participants in AoU, BioVU, and UKB, respectively.

### CHIP is associated with increased risk of incident SPRA, but not SNRA

In a three-cohort meta-analysis, a subtype-specific relationship between CHIP and risk of incident RA was observed, with a clear distinction between SPRA and SNRA. CHIP was associated with increased risk of incident SPRA (pooled hazard ratio (HR) 1.29; 95% CI 1.05–1.57; *p* = 1.3×10^-2^; **Fig. 2)**. Gene-stratified analyses revealed that this association was primarily driven by *DNMT3A*-CHIP (HR 1.42; 95% CI 1.10–1.82; *p* = 7.1×10^-3^). By contrast, *TET2*-CHIP was not associated with incident SPRA (HR 0.99; 95% CI 0.63–1.56), nor was *ASXL1*-CHIP (HR 1.51; 95% CI 0.78–2.92; **Fig. 2**). Neither overall CHIP nor any specific CHIP driver gene was associated with incident SNRA (**Fig. S3**). The distribution of CHIP driver mutations was similar between individuals who developed SPRA and those who developed SNRA **(Fig. S4)**.

**Figure 2.**
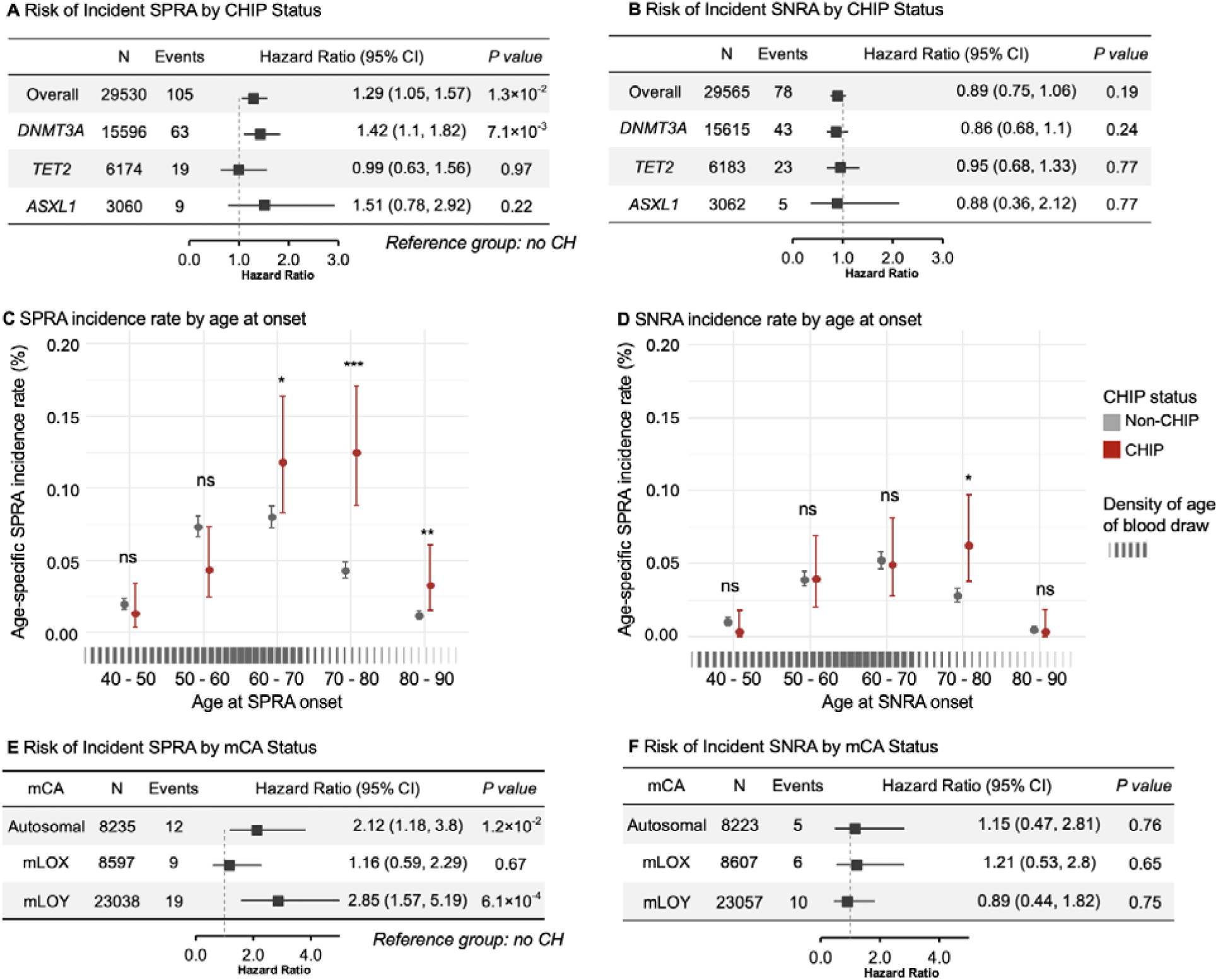
Association of clonal hematopoiesis with incident SPRA and SNRA. Forest plots show meta-analytic association of CHIP and specific CHIP driver genes (*DNMT3A, TET2, ASXL1*) with SPRA **(A)** and SNRA **(B)** incidence. CHIP is associated with an increased risk of incident SPRA, driven primarily by *DNMT3A* CHIP. Autosomal mCA and mLOY are associated with an increased risk of SPRA. No somatic genetic factors were associated with statistically significant effect on risk of SNRA. The column “N” indicates the number of CH carriers included, and “Events” indicate the number of incident SPRA/SNRA cases among CH carriers. Points indicate estimated hazard ratio (HRs) and horizontal lines indicate 95% confidence intervals. Age-specific incidence rates of SPRA **(C)** and SNRA **(D)** incidence were calculated across categories of RA onset age and compared between individuals with CHIP (red) and without CHIP (gray). Error bars represent 95% confidence intervals. A single biobank (UKB) shows the associations of mCA (autosomal mCA, mLOX, mLOY) with SPRA **(E)** and SNRA **(F)** incidence.

### Autosomal mosaic chromosomal alterations and mosaic loss of the Y chromosome are associated with increased risk of incident SPRA, but not SNRA

Survival analyses in UKB (the only biobank where mCA data is available), revealed striking associations with SPRA but none for SNRA. Autosomal mCA was associated with an increased risk of incident SPRA (HR 2.12; 95% CI 1.18–3.80; *p* = 1.2×10D^2^). Among male participants, mLOY was associated with an increased risk of SPRA (HR 2.85; 95% CI 1.57–5.19; *p* = 6.1×10D^4^). No significant association was observed between mLOX and risk of SPRA among female participants (HR 1.16; 95% CI 0.59–2.29), nor between any class of mCA and incident SNRA.

### CHIP^+^ SPRA is no more inflammatory than CHIP^-^ SPRA

Given the association between CHIP and incident SPRA, we sought to assess whether participants with CHIP developed a sub-type of SPRA with higher levels of systemic inflammation as manifested in the C-reactive protein level. Therefore, we compared the last CRP measurement prior to diagnosis of incident SPRA between patients with/without CHIP and compared to controls without CHIP who developed osteoarthritis (OA). SPRA groups exhibited substantially higher CRP levels than OA controls (*P* < .001). CRP levels in the CHIP^+^ SPRA group were numerically higher than those in CHIP^-^SPRA (median 9.1 vs. 8.5 mg/L) but the difference was not statistically significant (*p* = 0.06; **Fig. S5**).

### CH is not associated with prevalent RA

In contrast to the clear associations between CH and incident SPRA, there was no association between CH and prevalent RA, neither SPRA nor SNRA. (**Fig. S6**). In meta-analysis across the three cohorts, the odds of having prevalent SPRA did not differ between CHIP carriers and non-carriers (OR 0.93; 95% CI 0.81–1.07, *P* = 0.31), nor were *DNMT3A*-, *TET2*-, or *ASXL1*-CHIP associated with increased SPRA prevalence. Similarly, in the UK Biobank autosomal mCA was not enriched among people with prevalent SPRA (OR 1.41; 95% CI 0.99–2.02), nor was mLOX (OR 0.94; 95% CI 0.64–1.37) or mLOY (OR 1.26; 95% CI 0.91–1.75). Similar patterns were observed for prevalent SNRA, with no significant associations for overall CHIP, individual driver genes, or any mCA type.

### CHIP is associated with late-onset SPRA but not early-onset SPRA

The association of CHIP with incident but not prevalent SPRA presented an apparent conflict. We investigated further by examining the incidence of SPRA by decade and comparing to the age when blood samples were taken for DNA sequencing **(Figure 2C, 2D)**. Most blood samples were taken in middle age and CHIP carriers exceeded non-carriers in SPRA incidence only in older age. The largest CHIP-associated difference in SPRA incidence was observed among research participants age 70-80, an RA subtype that has been termed late-onset RA.^18^ In contrast, there was no difference in SPRA incidence by CHIP status among people with age < 60, suggesting that early-onset RA is largely unrelated to CHIP.

### CHIP and all-cause mortality among patients with SPRA and SNRA

Differential mortality is of absolute interest and could also explain the apparently conflict described above, therefore we examined the risk of all-cause mortality among research participants with prevalent SPRA and SNRA with/without CHIP. Among individuals with SPRA, CHIP carriers had shorter survival than non-carriers by the log-rank test (*p* = 0.04; **Fig. 3A**). Age-scale survival analysis revealed that overall CHIP was not significantly associated with mortality (HR 1.39; 95% CI 0.92–2.09), but *TET2*-CHIP was associated with a more than twofold increased risk of mortality (HR 2.36; 95% CI 1.11–5.03; *p* = 2.6×10^-2^; **Fig. 3B)**. Analysis of other common CHIP drivers and mCA among participants with SPRA found no association with mortality (**Fig. 3B**). In contrast to SPRA, neither CHIP nor any sub-type nor any mCA was associated with mortality among participants with SNRA.

**Figure 3.**
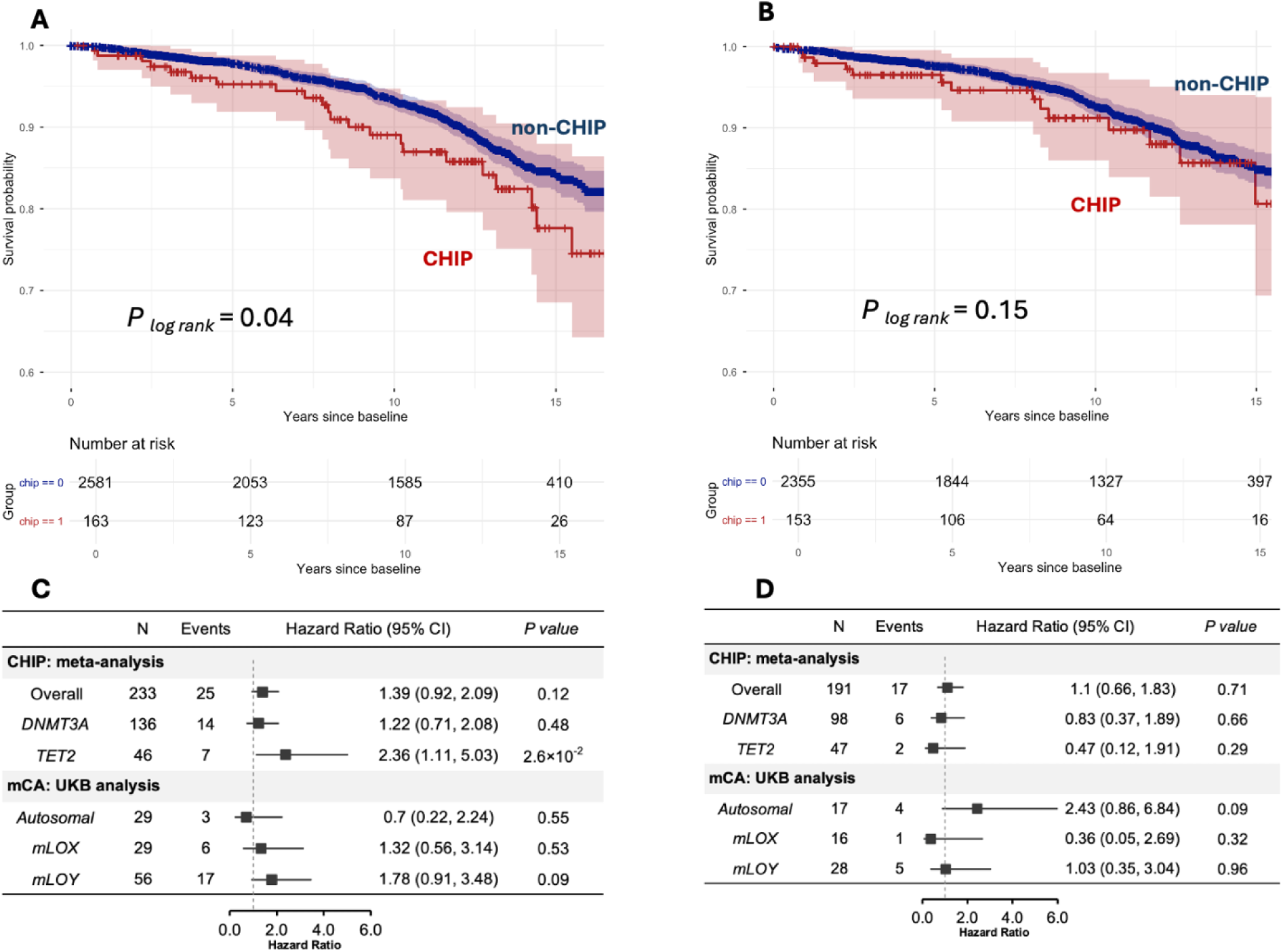
The association between CH and mortality among participants with RA. Panels A and B display Kaplan-Meier estimates of overall survival for patients with prevalent SPRA (**A**) and SNRA (**B**), stratified by CHIP status. Shaded areas represent 95% confidence intervals, and number at risk are shown below each plot. Age-scale Cox proportional hazards models were performed to evaluate the association between overall CH and specific CH driver genes with all-cause mortality among SPRA (**C**) and SNRA (**D**) patients.

### Genetically predicted C1s and C1r modify the effect of CHIP on risk of SPRA

To investigate the role of classical complement in mediating the CHIP-associated risk for SPRA, we performed age-scale survival analysis with an interactive effect between genetically predicted complement levels and CH status. These analyses revealed effect modification between CHIP and C1s (*p* = 1.0×10^-2^, *p*_adj_ = 0.049) and C1r (*p* = 1.6×10^-2^, *p*_adj_ = 0.049), indicating that higher genetically predicted complement levels attenuate the CHIP-associated risk of SPRA. In contrast, no significant interaction effect was observed for SPRA and any other complement components (*p* > 0.2; **Figure 4**), nor any complement components with SNRA. A sensitivity analysis conducted only among people with CHIP examined the effect of genetically predicted complement levels on risk of SPRA and observed consistent effects (C1s: OR 0.80; 95% CI 0.68–0.94, *P* = 7.8×10^-3^; C1r: OR 0.78; 95% CI 0.65–0.93, *P* = 6.7×10^-3^) **(Fig. S7)**.

**Figure 4.**
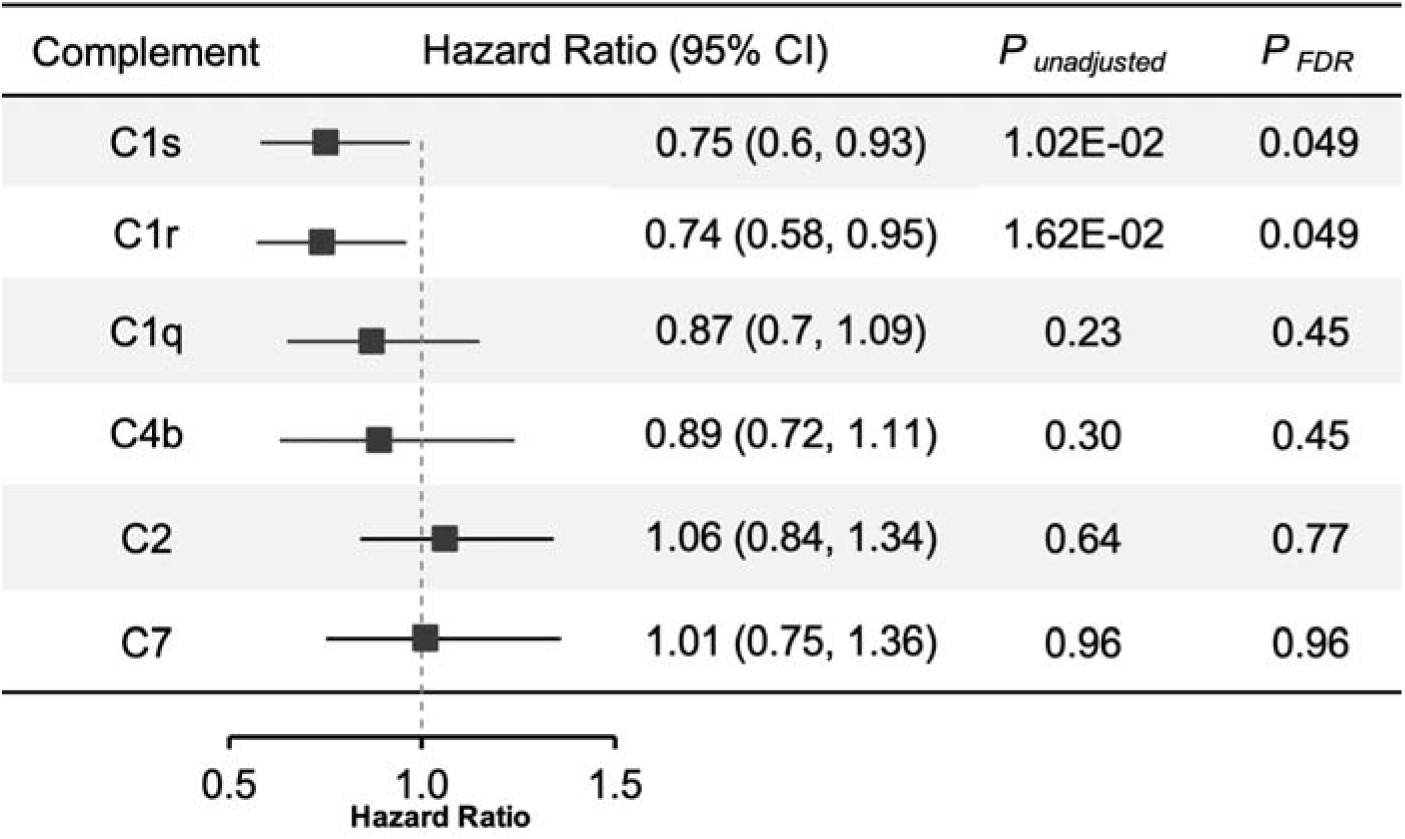
Effect modification of CHIP-SPRA association by genetically predicted complement levels. Forest plots show hazard ratios (HRs) and 95% confidence intervals for the interaction between CHIP and genetically predicted complement components on the risk of incident SPRA. Interaction terms were evaluated using age-scale Cox proportional hazards models to account for the time-to-event structure of incident SPRA. Effect modification was observed for genetically predicted C1s and C1r, with higher levels attenuating the association between CHIP and SPRA risk, whereas no significant interactions were detected for other complement components. *P* values are shown before (*P*_unadjusted_) and after false discovery rate (FDR) correction (*P*_FDR_ also known as *q*). Points and lines represent hazard ratios and 95% confidence intervals.

## Discussion

Our three-biobank meta-analysis found that CHIP was associated with an increased risk of incident SPRA, driven primarily by an increase in *DNMT3A*-CHIP-associated late-onset SPRA. Furthermore, participants with *TET2*-CHIP and SPRA had an increased risk of all-cause mortality compared to participants with SPRA alone. Effect modification analysis found genetically predicted levels of classical complement proteins C1r and C1s may modify the CHIP associated risk of SPRA. In UKB, autosomal mCA and mLOY were associated with an increased risk of SPRA.

The association between CHIP and incident SPRA with absence of association between CHIP and prevalent SPRA presents an apparent conflict. Straightforwardly, this observation may be the result of the reality that CHIP rarely occurs before middle age and RA generally onsets in older adulthood. However, results presented here suggest that differential survival may explain part of the enigma, with CHIP^+^ patients with SPRA less likely to survive to the date of enrollment. Intriguingly, a final possibility is that the treatments rendered for RA may cause CHIP clones to shrink or resolve, as has been observed experimentally for one RA treatment, tocilizumab.^32^

Throughout this study all analyses relating to SNRA were null. With 1,310 cases of incident SNRA, power was adequate to detect an effect if it were present. The discordant results between SPRA and SNRA could be due to biological differences between SPRA and SNRA or, as SNRA is a clinical diagnosis without confirmatory objective data, they could reflect the difficulty of studying SNRA with a biobank approach due to the high susceptibility for misclassification.

The strong association between mLOY and incident SPRA is consistent with prior work on prevalent SPRA.^18^ Still, it was observed only in one biobank and merits replication in other biobanks. Similarly, the association between autosomal mCA and SPRA was statistically sound but merits replication. If these effects were found to be consistent across cohorts, mechanistic studies could begin to determine the pathways that relate these larger scale somatic genetic changes to RA.

The strengths of this study include its large sample size, considering >1 million research participants for inclusion/exclusion and ultimately studying 612,989. The large sample size permitted use of a phenotyping approach that prioritized specificity over sensitivity, as is critical for valid inference of uncommon outcomes. The phenotyping approach combined diagnosis codes, laboratory values, and medication prescriptions, yielding an estimated 99% specificity and overcoming a critical weakness of recent biobank-based work on CH and RA which ascertained RA status strictly from billing codes.^14,15,20,33^ This study also made use of the longitudinal follow-up data in each biobank to conduct age-scale survival analysis, suggesting that CH causes RA, rather than *vice versa*.

This study also has notable limitations. First, because general-purpose genetic sequencing was used to infer CHIP status, the ascertainment was neither sensitive nor specific.^21^ Imperfect CHIP calls increase the risk of false negative studies, and attenuate the discernable risk, but do not increase the risk of false positive associations.^21^ Furthermore, the size of a CHIP clone can have profound implications for health outcomes, but cannot be estimated with general purpose sequencing.^11,34^ To address these weaknesses will require dedicated CHIP sequencing in the baseline samples of a cohort that has been followed longitudinally.

Second, the heterogeneity of biobank designs might complicate direct comparison of results across cohorts. AoU is a population sample where participants provide a blood sample at a time of their choosing, while BioVU is a hospital-based cohort where blood samples left over after clinical testing were used for sequencing, and UKB is a prospectively designed study where participants were invited to the assessment center on a medically arbitrary. To address this heterogeneity of baseline, we conducted survival analyses on the age-scale rather than on the time scale.^35^ Furthermore, we examined heterogeneity across cohorts using the I² statistic. We observed low heterogeneity for the overall analysis and for *DNMT3A*-CHIP analyses (I^2^ <35%), supporting the robustness of the study’s central findings.

Third, because incident SPRA and SNRA are uncommon and our approach prioritized specificity over sensitivity, there were few events among participants with each kind of CH. A cohort in which more participants had CH or more participants developed RA would provider more powerful statistical inference.

In summary, SPRA has long been known to be a complex auto-immune disease with antigen-driven clonal proliferation of lymphocytes as a core pathophysiologic process. The results presented here suggest that the pro-inflammatory, antigen-independent clonal proliferation of hematopoietic stem cells also plays a role in SPRA pathogenesis. The classical complement cascade emerged as a promising pathway that may mediate the relationship. Future work into mechanisms by which *DNMT3A*-CHIP, mCA, and mLOY drive this increased risk of SPRA merit further study as these insights might guide development of early interventions for patients with CHIP to avoid SPRA or precision interventions for patients with SPRA based on their CHIP status.

## Supporting information

supplemental ables

## Data Availability

All data produced in the present study are available upon reasonable request to the authors except those owned by the UKBiobank, NIH AllOfUs, and Vanderbilt BioVU consortia, which each have their own access protocols.

https://www.ukbiobank.ac.uk/

https://allofus.nih.gov/

https://www.vumc.org/dbmi/biovu

## Acknowledgments and affiliations

This work was supported by the NIH DP5OD029586, the Burroughs Wellcome Fund, the RUNX1 Research Program, the Pew Charitable Trusts (AGB) and the Arthritis National Research Foundation 128808 and Rheumatology Research Foundation (RWC).

## Key messages

**What is already known on this topic**: Clonal hematopoiesis (CH) is an asymptomatic condition of aging associated with risk for many inflammatory diseases of aging. Recent cross-sectional studies suggest an association with rheumatoid arthritis (RA), but lacked longitudinal follow-up and phenotype specificity.

**What this study adds**: This multi-ancestry study of >600,000 research participants used rigorous phenotyping and longitudinal clinical follow-up to discover that CHIP, especially when drive by mutation in *DNMT3A* is associated with an increased risk for seropositive RA. Genetic follow-up nominates the classical complement cascade as a potential mediator of the relationship.

**How might this study affect research, practice or policy**: Future research should investigate whether specific therapies could abrogate the CHIP-associated risk of SPRA and whether CHIP+ patients with RA may merit a personalized management approach.

---

**Figure S1.**
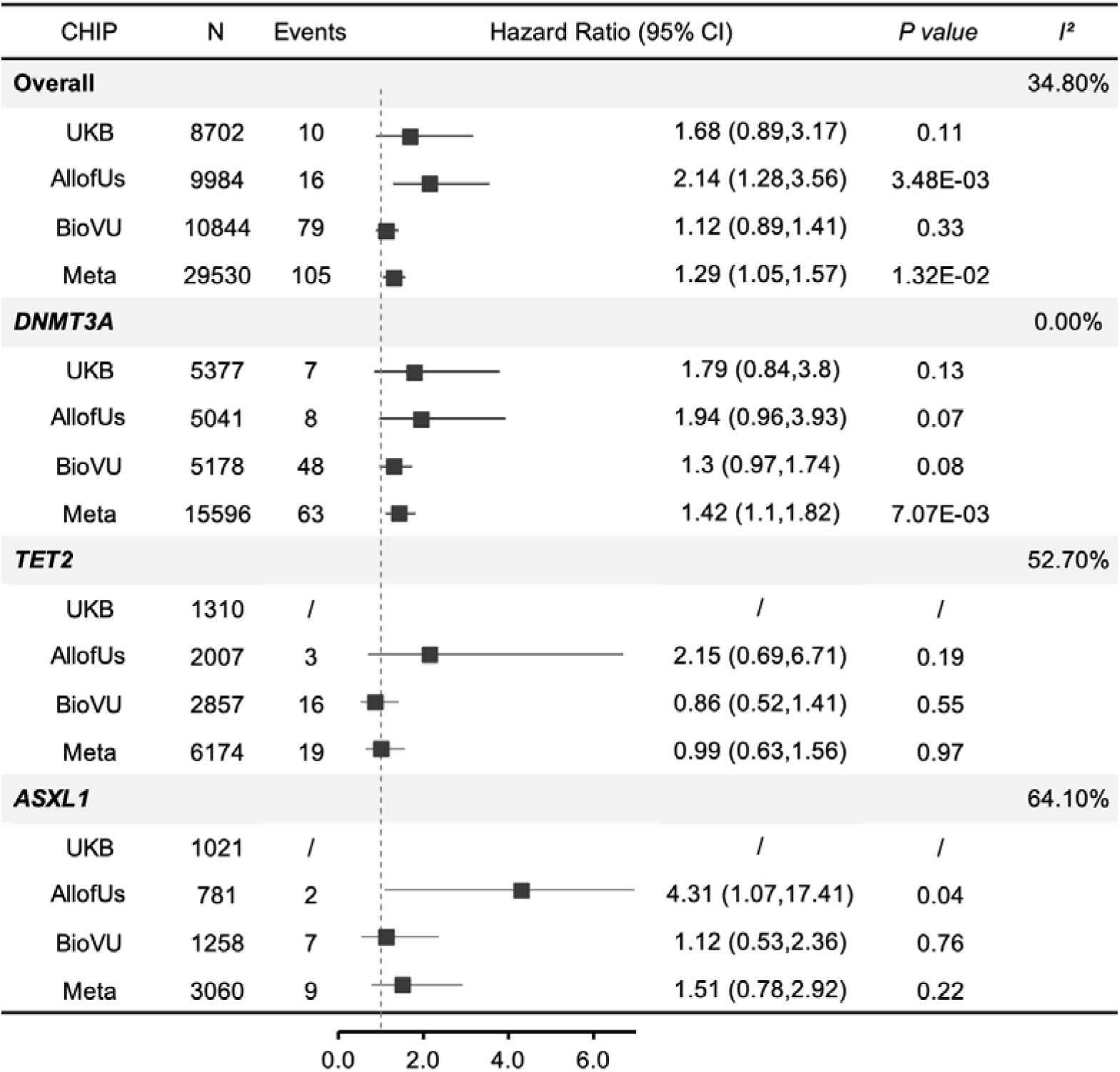
Time-to-event association of CHIP with incident SPRA by cohort. This forest plot shows the associations of overall CHIP and individual driver gene mutations (*DNMT3A, TET2, ASXL1*) with SPRA incidence in each cohort and after meta-analysis. The column “N” indicates the number of CH carriers included, and “Events” represents the number of RA cases among these CH carriers. Hazard ratios (HRs) with 95% confidence intervals quantify the effect sizes.

**Figure S2.**
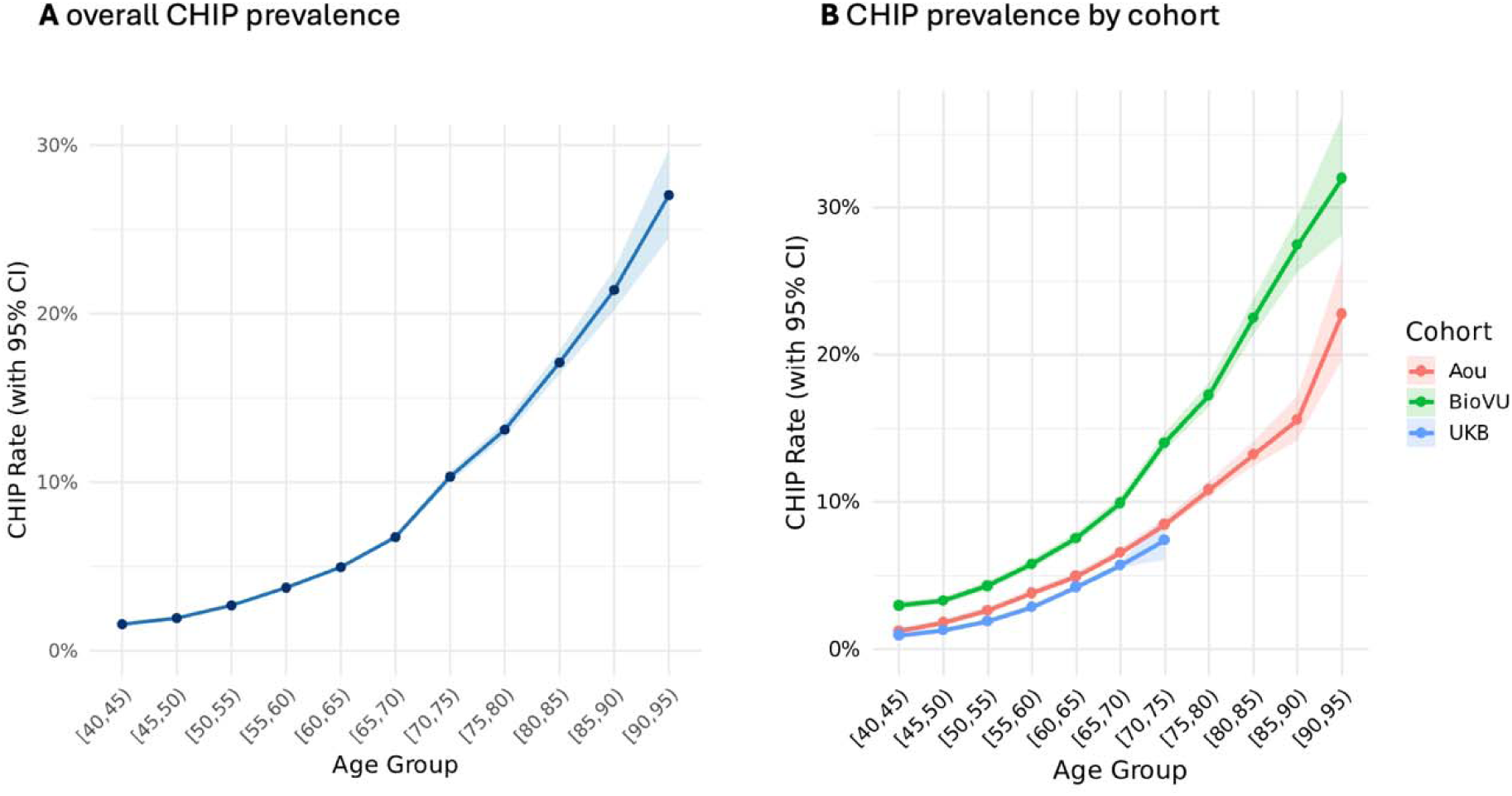
Age-related prevalence of CHIP across cohorts. (**A**) Overall prevalence of CHIP increased steadily with advancing age, exceeding 10% after age 70. (**B**) Age-stratified CHIP prevalence across the BioVU, *All of Us*, and UK Biobank cohorts, showing consistent age-dependent increases but higher absolute rates in BioVU.

**Figure S3.**
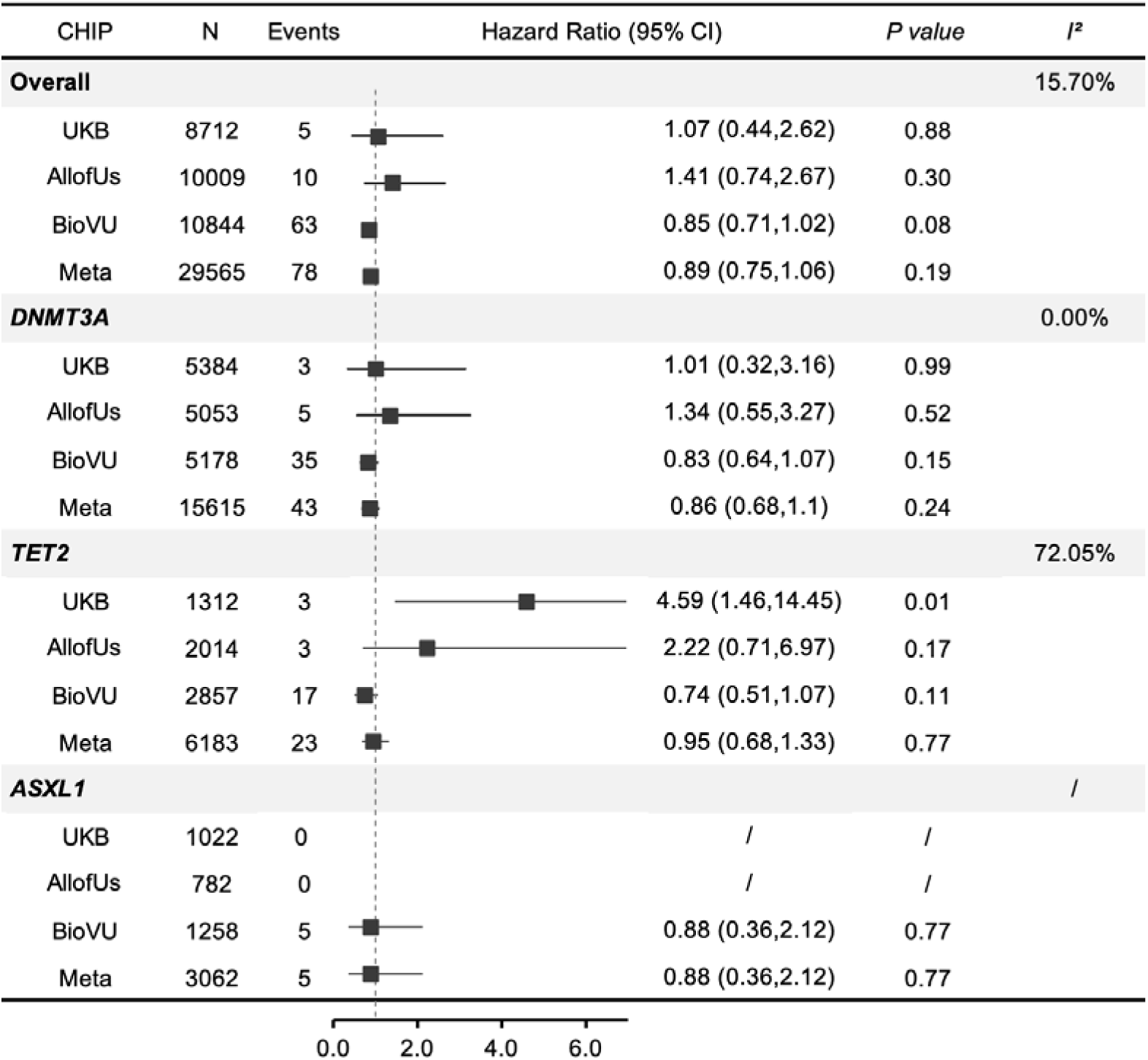
Time-to-event association of CHIP with incident SNRA by cohort. This forest plot shows the associations of overall CHIP and individual driver gene mutations (*DNMT3A, TET2, ASXL1*) with SNRA incidence in each cohort and after meta-analysis. The column “N” indicates the number of CH carriers included, and “Events” represents the number of RA cases among these CH carriers. Hazard ratios (HRs) with 95% confidence intervals quantify the effect sizes.

**Figure S4.**
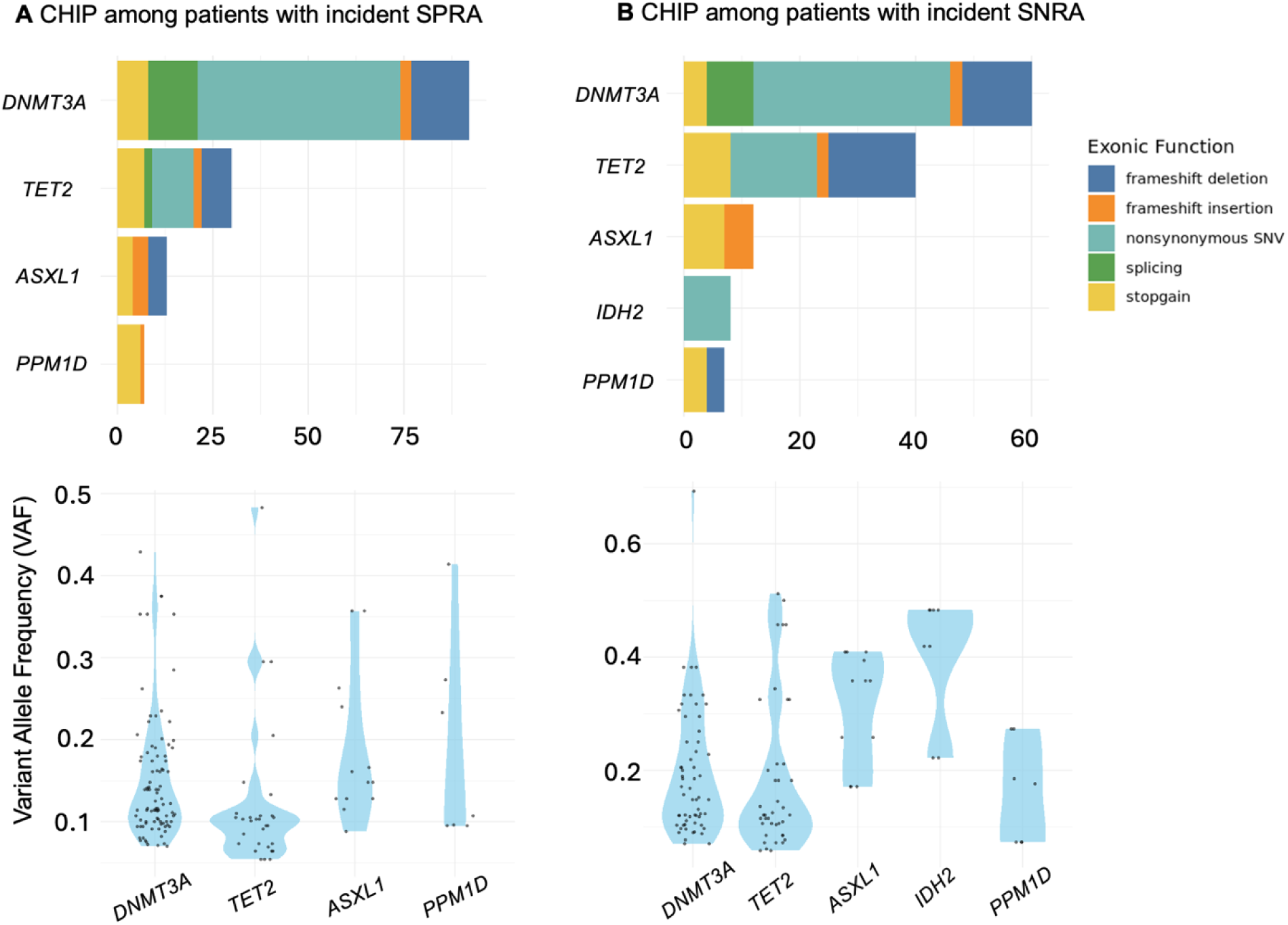
CHIP driver gene distribution among SPRA and SNRA patients. Panels A and B display the distribution of CHIP driver mutations among individuals who developed seropositive RA (SPRA; left) and seronegative RA (SNRA; right). Bars represent the number of carriers for each gene, stratified by exonic function (frameshift deletion, frameshift insertion, nonsynonymous SNV, splicing, and stop-gain mutations). The bottom panels show the variant allele frequency (VAF) distributions for each driver gene among patients with incident SPRA (left) and SNRA (right). Violin plots depict the range and density of clone sizes, with individual data points overlaid.

**Figure S5.**
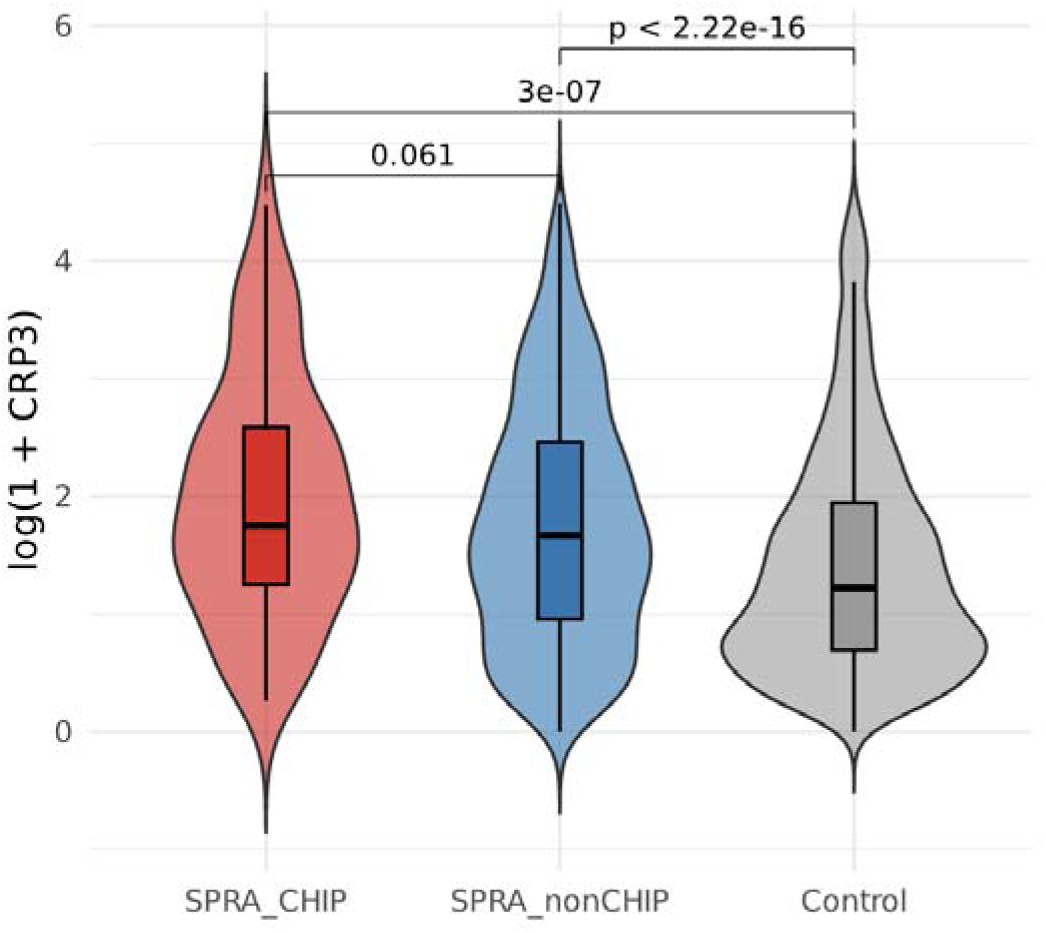
CRP levels among SPRA patients with and without CHIP and osteoarthritis controls. Violin and box plots show the distribution of log-transformed CRP levels in three groups: SPRA patients with CHIP (SPRA_CHIP), SPRA patients without CHIP (SPRA_nonCHIP), and osteoarthritis controls. Pairwise comparisons were performed using Wilcoxon rank-sum tests with continuity correction. Boxplots depict medians and interquartile ranges, and violins represent kernel density estimates.

**Figure S6.**
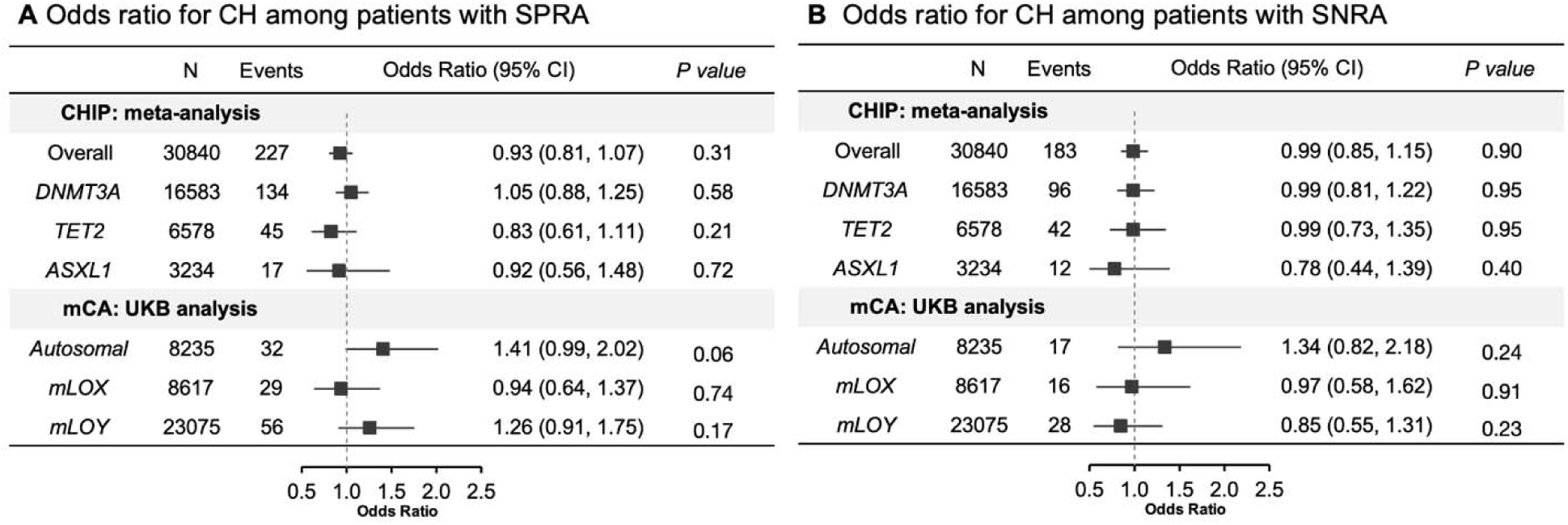
Association of clonal hematopoiesis with RA prevalence. This forest plot shows the associations of overall CHIP and individual driver gene mutations (*DNMT3A, TET2, ASXL1*) with SPRA (**A**) and SNRA (**B**) prevalence after meta-analysis, and the associations of mCA (autosomal mCA, mLOX, mLOY) with RA prevalence in UKB. Logistic regressions were performed to test the associations after adjusting the same covariates as mentioned in Methods. The column “N” indicates the number of CH carriers included, and “Events” represents the number of RA cases among these CH carriers. Odds ratios (ORs) with 95% confidence intervals quantify the effect sizes.

**Figure S7.**
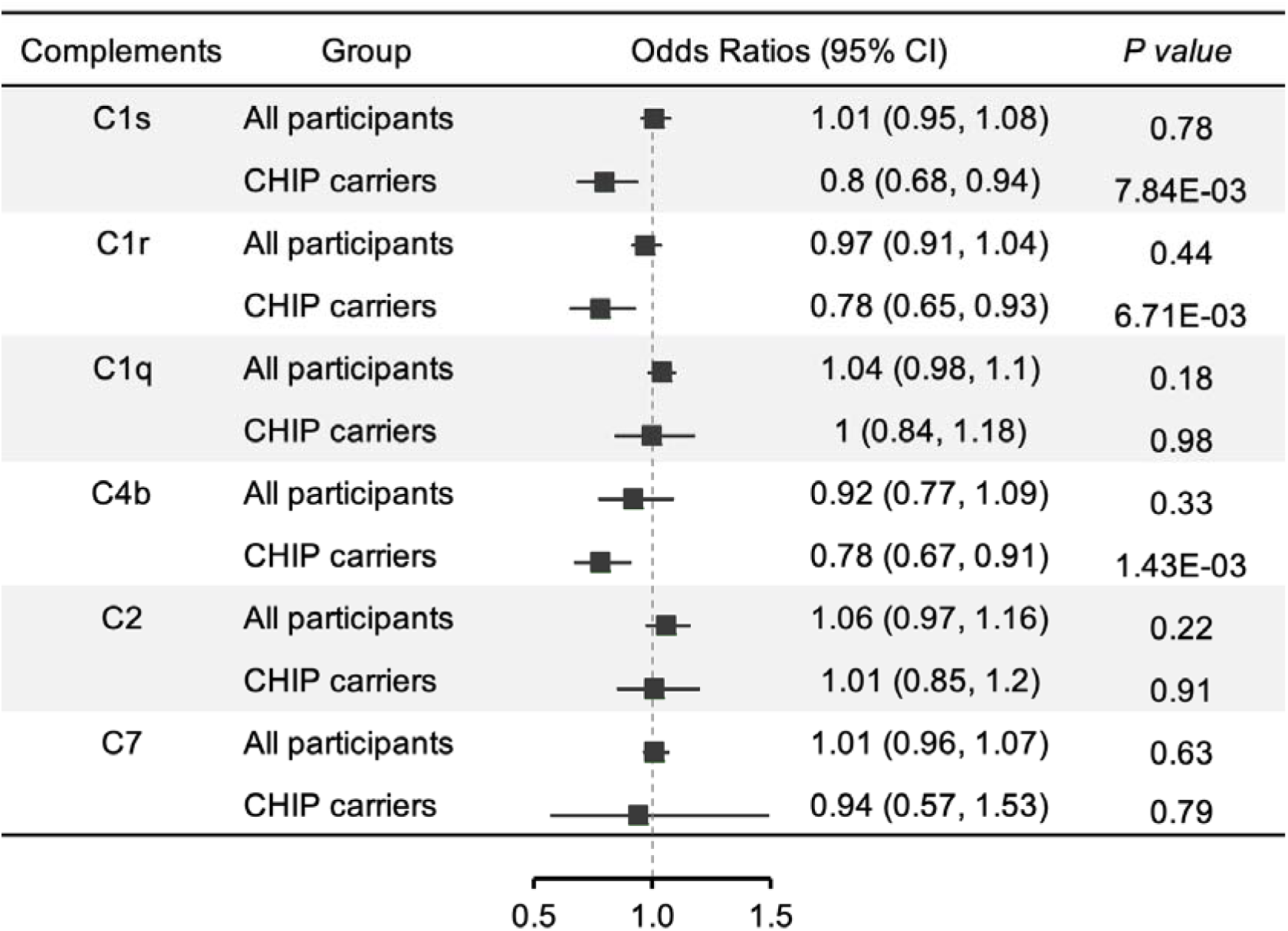
Associations of genetically predicted Complements with SPRA risk in the overall population and among CHIP carriers. Forest plots show odds ratios and 95% confidence intervals for the associations between genetically predicted complement components and SPRA risk in all participants and when restricted to individuals carrying CHIP. Because genetically predicted complement levels reflect stable, lifelong genetic predisposition rather than time-varying exposure, their associations with SPRA risk in the overall population and among CHIP carriers were evaluated using logistic regression models. Odds ratios represent the change in SPRA risk per unit increase in genetically predicted complement level. While no significant associations were observed in the overall population, genetically predicted C1s, C1r, and C4b were associated with lower odds of SPRA among CHIP carriers.

