## supplemental appendix for "Clonal Hematopoiesis is Associated with Risk of Incident, Seropositive Rheumatoid Arthritis"

Supplementary appendix

Diagnosis codes

In the UKB, relevant codes were identified by searching for concepts that match the regular expression ‘(R|r)heumatoid’ but not ‘(J|j)uvenile’, yielding 130 codes, which were manually reviewed to ensure no inappropriate codes were captured. RA SNOMED codes were categorized as SPRA, SNRA, or RA NOS based on matching either “positive” (17), “negative” (6), or neither (107) (**Supplementary Table 2**). Diagnosis codes were evaluated with no consideration as to whether they were assigned inpatient vs. outpatient.

Laboratory values

We identified concepts that capture rheumatoid factor (RF) and anti-cyclic citrullinated peptide (CCP) by searching the concept table, subsetted to measurement concepts, and manually reviewing resulting concepts. For RF we searched for concepts that match ‘(R|r)heumatoid factor’. For CCP, we searched for concepts that match ‘citrullinated’ but not ‘Homo|Urine|vimentin|viral’.

Quantitative laboratory values that had an associated upper limit of normal (ULN) were adjudicated as positive or negative based on comparison with the ULN. Quantitative laboratory values in AOU and BioVU without defined ULN were adjudicated based on the ULN present in other measurements with the same units, with manual review to ensure at least 90% of results were negative and consistency with clinical experience of the authorship team in the practice of rheumatology in the United States. This process yielded a cutoff for RF of 15 IU/mL, for CCP in U/mL of 20 U/mL and for unitless CCP of 0.94. Quantitative lab values in UKB without defined ULN were adjudicated based on the same quantile analysis and a review of the UK’s National Health Services Pathology Lab publications, which yielded a cutoff of 20 IU/mL for RF and 7 U/mL for CCP. Laboratory values that were textual in nature were considered positive if they matched ‘pos’ or ‘>’, negative if they matched ‘neg’ or ‘<’, and N/A otherwise.
